# Genetic Connection to Drug-Induced Liver Injury (DILI) Through Statistical Learning Methods

**DOI:** 10.1101/2021.02.15.21251747

**Authors:** Roland Moore, Kristin McEuen

**Affiliations:** Florida State University; US Government

**Keywords:** DILI, SNP, Genomic, Interactions, Associated, Epistasis, Splines, bilirubin

## Abstract

Drug-Induced Liver Injury (DILI) is one of the major causes of drug development failure or drug withdrawal from the market after development. Therefore, investigating factors associated with DILI is of paramount importance. Environmental factors that contribute to DILI have been investigated and are, by and large, known. However, recent genomic studies have indicated that genetic diversity can lead to inter-individual differences in drug response. Consequently, it has become necessary to also investigate how genetic factors contribute to the development of DILI in the presence of environmental factors. Thus, our aim is to find appropriate statistical methods to investigate gene-gene and/or gene-environment interactions that are associated with DILI. This is an initial study that only explores statistical learning methods to find gen-gene interactions (epistasis). We introduce Multifactor Dimensionality Reduction (MDR), Random Forest (plus logistic regression), and Multivariate Adaptive Regression Splines (MARS), as the few potential methodological approaches that we found. Next, we attempt to improve the MARS method by combining it with a variable selection method.

## 1 Introduction

Drug Induced Liver Injury (DILI) is the damage to the liver as a result of a taking a drug. The human liver is vulnerable to drug injury because most drugs are assimilated into the body through the liver. Thus, if the drug is toxic, it causes damage/injury to the liver. Hence, the result is DILI. In most drug development clinical trials, DILI is one of the main valid causes for bringing the entire development process to a halt. Even after the drug has been successfully developed, because of DILI, it can be recalled. Moreover, even when a drug is deemed safe enough and released to the market, some relatively small fraction of the population that take the drug can get DILI [3]. Therefore, the US government funds research to identify the factors that promote DILI in order to find solution for preventing, reducing and/or curing DILI.

### 1.1 Drug-Host-Genetic Factor Interplay

Non-genetic factors that contribute to the risk of having DILI has been well researched already: Chen et al (2015) [3] established that host factors like age, disease, etc., and drug properties like daily dose, reactive metabolite, etc., and the interactions between these factors influence DILI risk [3]. However, how genetic factors contribute to DILI is not yet well established: According to Kaplowitz (2004) [13], there has not been any known methodology for investigating genetic causes of DILI. We therefore, intend to use statistical learning methods to find how drug-host-genetic factors interplay influence DILI risk, as depicted in the diagram below (figure 1). Thus, our research goals are to find the following:

**Figure 1:**
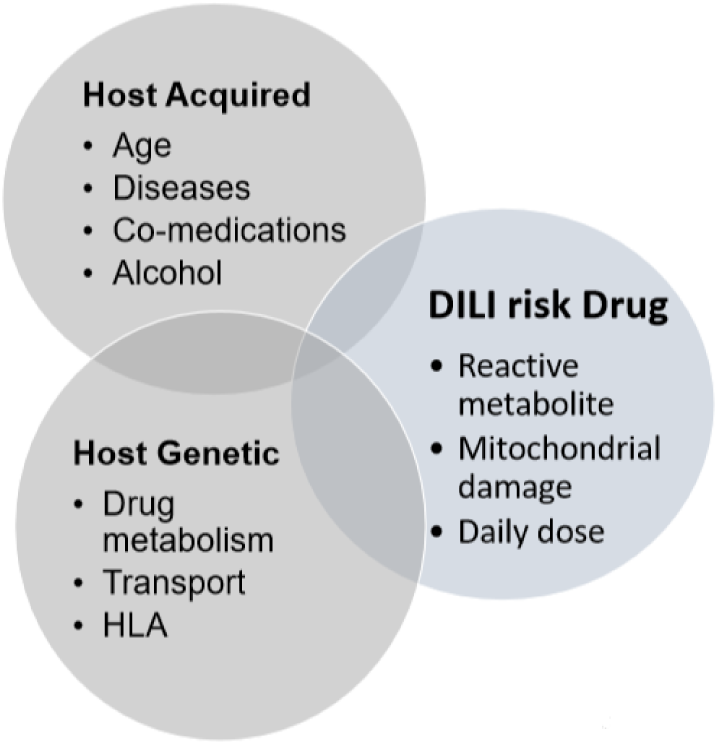
Drug-Host-Genetic Factor Interplay

1. Appropriate statistical learning methods to analyze data with genetic components in the form of SNPs.
2. Specific genes and environmental factors as well as their possible interactions that are associated with DILI.

### 1.2 Genetics Link

Genetic connection to DILI risk was motivated by the study done by Daly et al (2009) [5]: They found the gene HLA-B*5701 to be a risk factor: Thus, people who carry this allele have 80 times more frequency to develop DILI than those who do not. Therefore, the question we seek to answer is: What genes, in the presence of environmental factors with all possible rational interactions, are associated with DILI?

Genetic data comes in the form of single nucleotide polymorphism (SNP). Per the image in figure 2, SNP is the difference in single nucleotide positions in the chromosome. This positional difference of the nucleotide occur in about one percent of the human genome and is known to be responsible for about 90 percent of individual human differences [10]. Also, SNP does not change from generation to generation. Thus, SNP is responsible for the common genetic variations among people [10]. For example, SNP accounts for inter-individual differences in drug response. Therefore, genetics has become very important to the study of human health. Consequently, this has led to the recent increase in the study of personalized medicine [10].

**Figure 2:**
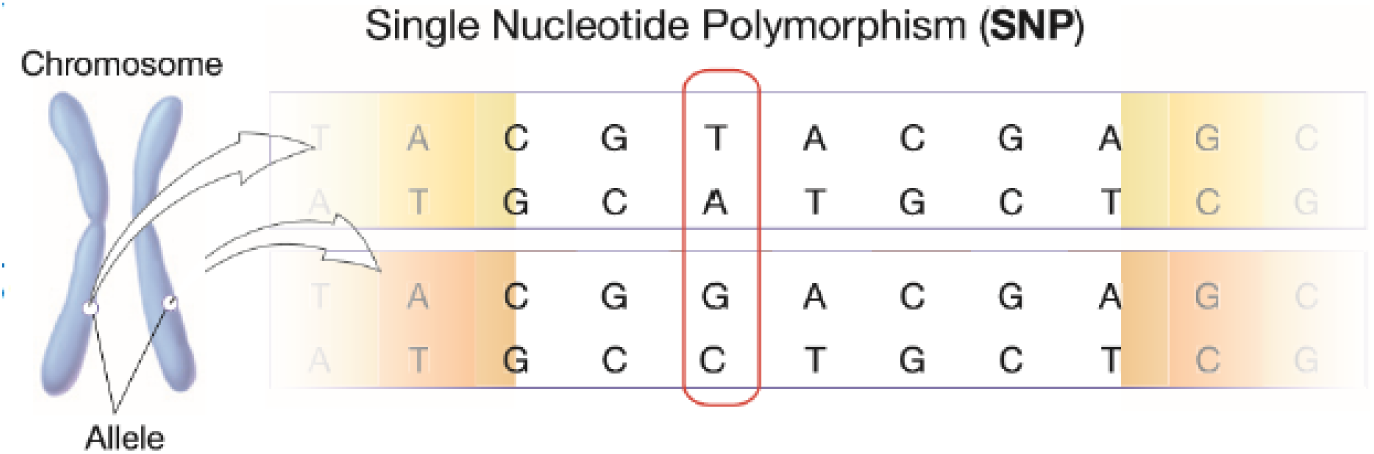
Displaying SNP- Sources: US National Library of Medicine

### 1.3 Role of Statistics

Statistical tools are necessary because statistics makes sense of randomness and variation. The statistical view is that the variability in the SNP or genes of a random individual accounts for such genetic factor contribution to the phenotype (DILI). Interactions involving genes (gene-gene and gene-environment) contribute significantly to complex human diseases and traits. Thus, one of the main sources of individual trait differences as well as variations in medical and drug responses could be these genetic interactions (Yi, 2010) [29]. Some statistical learning methods measure the interaction effects of certain explanatory variables on the response variable(s). This is further reason why we are using statistical methods for this study.

### 1.4 Findings From Literature Review

Statistical/machine learning methodologies for genetic contribution to diseases and other phenotypic traits through analysis of SNPs is relatively new and few. Literature also documents known challenges for SNP data analysis. I will outline some of the general and specific challenges to our study, and suggest ways to address them.

#### 1.4.1 Challenges

Firstly, the curse of dimensionality is a problem with statistical genetic data analysis. The reason being that sample size in SNPs data is relatively small compared to the number of SNPs. SNPs themselves come in huge numbers, sometimes hundreds of thousands in numbers. This makes SNP data very ‘wide’. Therefore, classical statistical methods like regression (and it’s various forms) are less feasible to be used for such high dimensional data. Secondly, some SNPs may be correlated with each other and therefore analysis can be complicated with correlated SNPs. Thirdly, because the SNP data size is huge in width, it is generally computationally intensive and time consuming. Finally, Statistical/machine learning methodologies needed for this particular project was relatively scarce. In some instances, the softwares and/or websites of some of the methodologies that we found were no longer in existence nor maintained.

### 1.5 Overall/Immediate Goal

The overall goal for this study is this: Find the statistical/machine learning methodologies that will identify drug-host-genetic factors and their interplay that influence DILI risk. However, we will immediately consider statistical/machine learning methodologies for only epistasis (statistical SNP interactions). Then, in the second stage of the study (will be a follow up to this paper), those discovered epistasis statistical/machine learning methodologies will be extended to the entire drug-host-genetic factors, to achieve the complete goal of the study.

## 2 Materials and Methods

The raw materials needed for this study are some general SNP dataset (for trials) plus a type of DILI SNP dataset (for actual testing). In addition, the statistical learning methods we chose were mainly informed by literature review on epistasis and gene-environment interactions articles. The statistical/ machine learning softwares used were mainly the MDR software package (http://sourceforge.net/projects/mdr/), and Salford Systems statistical with machine learning software (https://www.salford-systems.com/). Both software suits have graphical user interface. Hence, for the most part, coding was not required. In the next subsections I briefly describe the literature review information, selected statistical learning methods, and the data used in the study.

### 2.0.1 Addressing Challenges

Since the SNP data is wide (high dimension), Non-parametric procedures are usually preferable for this type of study. Sometimes, some machine learning methods combine non-parametric with a classical or parametric model such as logistic regression. Therefore, both parametric and non-parametric statistical methods are combined in some form, though most of the methodologies are non-parametric. Below is the brief outlines of the two main statistical models used in this project:

Parametric procedure outline; the Logistic Regression (Agresti, 2008) [1]:

If Y is the phenotype, DILI (binary; 1 or 0); *P* (*Y* = 1) = *p*, A is a SNP, B is another SNP, and AB is the interaction factor, then;

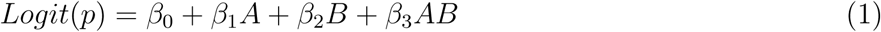

Non-parametric procedure outline:

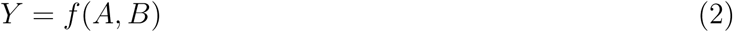

where the function f needs to be estimated directly from a particular data, using certain non-parametric function estimation procedure like Multivariate Adaptive Regression Splines(MARS) [6, 30, 27], Multifactor Dimensionality Reduction (MDR) [18, 19, 21], Random Forest [8], etc.

### 2.1 Epistasis Methods Found in Literature Review

Epistasis methods found in the review done by Wei et al. (2014) [25], Shang et al. (2011) [23], Lou et al. (2007) and other sources revealed the following methods (among many others) that are mentioned below without detail nor description: PLINK/ FastEpistasis [25], Multifactor Dimensionality Reduction (MDR) [21], SNPruler [25], Generalized Multifactor Dimensionality Reduction (GMDR) [16], SNP Harvester [25], Epistasis Detector Based on the Clustering of Relatively Frequent items, (EDCF) [25], Bayesian Epistasis Association Mapping (BEAM) [25], GenomeMatrix [25], Tree-Based Epistasis Association Mapping(TEAM) [25], Bayesian hierarchical Generalized Linear Model for haplotype interactions (BhGLM) [25], Random Forest (RF) [2], SNPInterForest [23], EPIMODE [23], and Multivariate Adaptive regression splines (MARS) [6].

### 2.2 Overview of the Three Methods Used

After thorough investigations, MDR, MARS and RF were chosen to investigate DILI epistasis. In the next subsections, brief descriptions of these three methods are given.

#### 2.2.1 Random Forest Plus Logistic Regression (RF)

##### Statistical context

- For a *p*-dimensional random vector *X*= (*X*_1_, …, *X*_*p*_)^*T*^ representing the data with predictor variables (SNPs), and a random binary variable *Y*, representing the phenotype (response), with an unknown joint distribution, *P*_*XY*_ (*X, Y*). We aim to predict *Y* from *f* (*X*).
- *f* (*X*) is determined by the loss function, *L*(*Y, f* (*X*)), and defined to minimize the expected value of the loss [*E*_*XY*_ (*L*(*Y, f* (*X*)))].
- The Zero-one loss function is a choice for measuring closeness of *f* (*x*) to *L* for the classification situation;

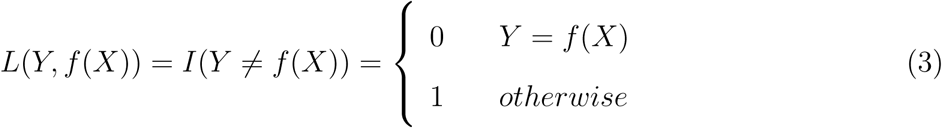
- Minimizing the *E*_*XY*_ (*L*(*Y, f* (*X*))) gives the Bayes Rule;

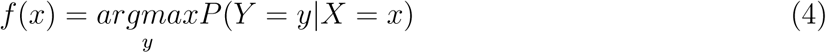
- Random Forest(RF) constructs *f* with a collection of base learning trees, *h*_*j*_(*x*), *j* = 1, …, *J*; and the predicted function is formed by majority voting as;

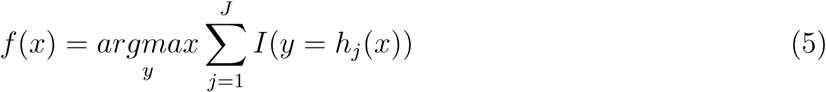

##### Brief description of algorithm

RF uses multiple decision trees as learning ensemble to give a single resultant output: Each tree acts on random bootstrap subsets of the data, as well as random subsets of the variables (SNPs) to make prediction. Each SNP (predictor) represents the node of each tree and a route links a sequence of predictor SNPs from the root to the leaves. The predictions from each of these trees are then aggregated in some way to give the overall output prediction-In classification, majority voting is employed. For further details on RF see Breiman (2001), Goldstein (2011), and Niel (2015) [2, 8, 20]. It must be pointed out that there is no actual model output from RF. Rather, RF gives a table of the ranking order of the important SNPs (variables) that contribute to the phenotype (DILI). Logistic regression is then used on the first few variables (say, first 10) to determine the interacting SNPs that are linked to DILI. The logistic regression part is subjective and not perfect. However, under the RF output structure, this is one of the best ways to proceed to the final result.

#### 2.2.2 Multifactor Dimensionality Reduction (MDR)

MDR was created primarily to detect gene-gene and/or gene-environment interactions in genetic data analysis. it is a non-parametric dimension reducing method. Here is a summary of the core algorithms [7]:

- Choose *d* factors (SNPs) with *l*_*i*_, *i* = 1, …, *d*, levels from *p* total factors.
- Selected factors (SNPs) are represented in *d*-dimensional space (contingency table) and in each cell, 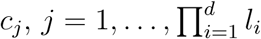, the case (*n*_1_) to control (*n*_0_) ratio

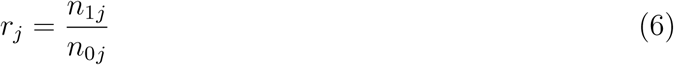

is calculated.
- *c*_*j*_ is labeled as high risk (H) if *r*_*j*_ > *T* (some threshold; e.g. *T* is equal to 1 for balanced datasets), as low risk (L) otherwise. Therefore, multi-locus genotypes are reduced into a one-dimensional variable.
- The core algorithm *k* models are evaluated by cross validation consistency (CVC), classification error (CE), and prediction error (PE) scores to choose the best model.
- CVC is the number of training sets in which a specific model has the lowest CE: The higher the CVC value, the better.

The following steps describes how it works:

**Step one**: To avoid over-fitting, the data is automatically divided into 10 equal parts, nine-tenths of which is used to train the data, and the last part is used for cross-validation to test the method’s accuracy. **Step two**: A selected subset of *d* genetic and/or environmental factors with their multi levels are represented in an *d*-dimensional space. Thus, for two loci, each with three genotypes (levels) interaction model, there will be nine two-locus-genotype possible combinations in a contingency table form. **Step three**: The ratio of the number of cases to the number of controls in each contingency table box is calculated. If this ratio is higher than pre-specified value (normally 1.0 for balanced data set), that particular interaction box is deemed as high risk for the phenotype (DILI). The model is then formed by combining all high risk interactions on one side and low risk ones on the other side. Therefore, the process with n dimensional space reduces to one dimension with two levels (high and low risk). **Step four**: The previous processes are repeated for all *d*-way interaction models, and the model with the least classification error is selected. **Step five**: Cross validation consistency (CVC), classification error (CE), and prediction error (PE) are used to assess the prediction prowess of the chosen model. The only difference between classification and prediction errors is that, the former is calculated using the training set while the latter uses the testing set. For further details on MDR, see Ritchie (2005), and Moore (2010, 2015) [21, 18, 19].

#### 2.2.3 MARS

The MARS Algorithm, through an automatic data set determined knots, divides the entire data set into many smaller regression subsets. Each of these subsets comes with a basis function, and all significant basis functions are aggregated to get the overall regression model. One clear advantage of MARS is that it gives a regression-like output model in the form *Y* = *BX* + *ϵ*; with the X’s being basis functions of the predictors here.

#### 2.2.4 MARS Overview

The MARS algorithm is such that the regression model is derived directly by the data, through a data driven automatic set of basis (hinge) functions with their corresponding coefficients. These basis functions are derived based on automatic data driven hinge values in the data, also referred to as knots and represented by the letter, *t*. As a particular linear relationship between the response and predictor variables in a data subset is being modeled, the knot automatically identifies the point of direction change from that relationship. This point of change now becomes a starting point for a new subset of another relationship; and the process continues with new knots and relationships to the end. Therefore, both linear and non-linear relationships with corresponding interactions are captured in the model.

#### 2.2.5 The MARS Model

MARS builds models of the following equation form;

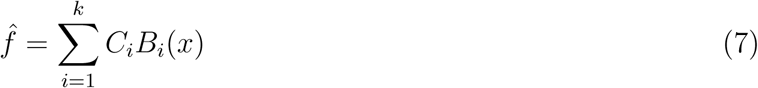

where each C is a constant coefficient multiplying a basis function B.

#### 2.2.6 How the MARS Model is Built

The MARS model is built using some form of forward and backward selection process in the following way: Forward selection combines all significant basis regression functions and their interactions. Backward selection prunes the result from the forward selection to avoid over fitting. This gives the optimized final model. During pruning, basis functions are eliminated from the over-fitted model one at a time based on some form of residual-sums-of-squares criterion called GCV (Generalized-Cross-Validation) score. This score ranges from zero to one. A score approaching one contributes optimally and a score approaching zero contributes almost none to the response. The following is the GCV equation:

For N observations, d independent number of bases functions (effective degrees of freedom), p as the penalty for adding a base function; the pruning formula is given by:

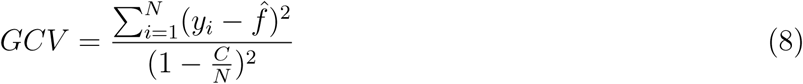

where *C* = 1 + *pd*

For more details on MARS, check out these references: Friedman (1991), Kane (2015), York (2001), and Yang (2004) [6, 12, 26, 30, 28]. Also, for a special reference with the Salford System software [22], check out https://www.salford-systems.com/products/mars.

## 3 Materials and Methods

### 3.1 Data Description

All datasets for this project are found here: https://ani.stat.fsu.edu/vic/DILIdata/. As shown in the table 1, data sets one and two are readily available simulated ones, retrieved from the MDR software package used for this project.

**Table 1:** Datasets for Genetic DILI

| Table 1: Datasets for Genetic DILI |  |  |
| --- | --- | --- |
| Data | Observation | SNP variables |
| Data Set 1 (Known SNPs) | 250 | 25 |
| Data Set 2 (Known SNPs) | 250 | 50 |
| DILI Chronicity Data | 271 | 872 |

The chronicity DILI data was generated by a genotyping method using microarray bead chip. It was acquired from International DILI Consortium, part of the International Serious Adverse Event Consortium (ISAEC); website; https://dataportal.saeconsortium.org/) [9]. There are 271 obervations (238 acute vs 33 chronic), and 872 SNPs (related to bile acid pathways). As further information, chronic DILI is relatively rare among DILI risk patients and doesn’t have a uniform definition. All the same, according to (Medina, 2016) [17], it can be thought of as occurring when abnormal serum chemistry values are measured (or have evidence of continued liver injury) for at least 6 months.

For this data set, DILI chronicity is dichotomized into the following: Chronic DILI is liver injury persisting for six months or more, coded as chronic (1); acute is liver injury occurring for less than 6 months, coded as acute (0).

## 4 Results

In these section and subsections, we outline the results of applying the selected statistical learning methods to the our SNP datasets.

### 4.1 Learning Methods Comparison

The three methods were compared using two available simulated data each with two known SNP Interactions. The recovery rate results for these interactions are displayed in the table 2. Recovery rate is the fraction of the known SNP interactions that are recovered by the method.

**Table 2:** Recovery rates for all the methods

| Recovery rate | MDR | RF + regression | MARS |
| --- | --- | --- | --- |
| Data set 1 (Known SNPs) | 100% | 100% | 100% |
| Data set 2 (Known SNPs) | 100% | 50% | 50% |

- Known SNPs are SNPs that were predetermined to be associated with the response variable (phenotype).
- Recovery Rate is the number of known SNPs that were recovered by the method.

For data set one, the recovery rates for all the three methods were perfect. However, for data set two, while the recovery rate for MDR was perfect for the known interacting SNPs, that of RF and MARS was only 50 percent. These two methods recovered only one of the known interacting SNPs.

### 4.2 A Previous Relevant Study on The DILI chronicity data

A previous study on the DILI chronicity data found **increased total serum bilirubin were associated with the drugs causing chronic liver injury**. See the figure 3. This information is important because it has a connection to our study’s findings that follow next.

**Figure 3:**
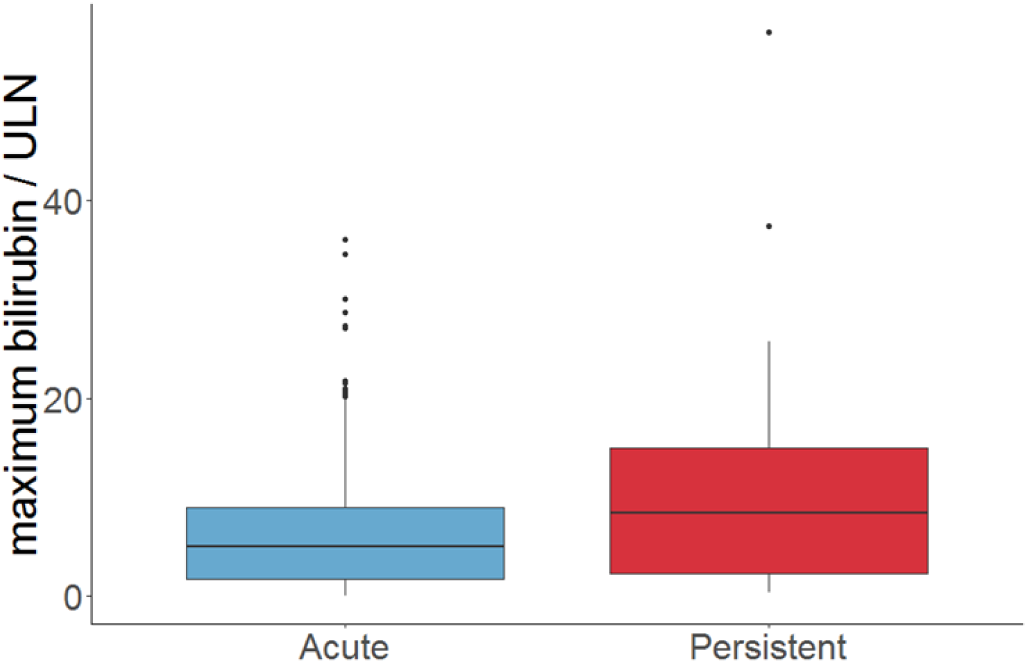
Chronic DILI-serum bilirubin connection

### 4.3 Epistasis from Chronicity DILI Data set

Per the results of epistasis in the table 3, each method’s SNP interactions have entirely different SNPs. It could be that though the SNPs are different, they may still share the same genetic biological information. However, we have not looked into that possibility of shared genetic biological information yet. We were however, able to identify the biological information on re6487213, an interacting gene, under the MARS model (further details in the next section).

**Table 3:** DILI Gene-gene interaction for all the methods

| Data | MDR | RF + Regression | MARS |
| --- | --- | --- | --- |
| DILI | rs7658048*rs5417 | rs5417*rs3785157 | rs6487213*rs3785157 |

**Table 4:** Preliminary Results For [rfe(RF)+ MARS] Method

| Recovery rate | MDR | RF + regression | MARS | rfe(RF)+MARS |
| --- | --- | --- | --- | --- |
| Data set 1 [25](Known SNPs) | 100% | 100% | 100% | 100% |
| Data set 2 [50] (Known SNPs) | 100% | 50% | 50% | 100% |

## 5 Discussion and Follow up Study

In the section, we will expatiate on the detected SNP biological information as well as introduce a follow up study we are working.

### 5.1 Detected SNP (rs6487213) Biological Info

The SNP, rs6487213, is a variant of the gene SLCO1B1, which is a solute carrier organic anion transporter family member 1B1: See https://ghr.nlm.nih.gov/gene/SLCO1B1. This gene is responsible for the instructions for synthesizing a type of protein called organic anion transporting polypeptide 1B1, or OATP1B1. This protein, found in liver cells, transports compounds from the blood into the liver to be eventually excreted from the body. Example of such a compound is bilirubin which is found in the bile within the liver. Other compounds transported are hormones, toxins and drugs.

Already, in a study of 9500 Caucasians, the allelic variation in SLCO1B1 was reported as a major genetic predictor of increased serum bilirubin levels [11]. Note that liver creates bile which contains bilirubin for food digestion. A healthy liver gets rid of the bilirubin while an unhealthy liver is unable to eliminate the bilirubin. So a higher amount of bilirubin presence in the blood indicate a liver disease, and hence a DILI occurrence.

### 5.2 Follow Up Study Preliminary Results

According to our findings, MARS was faster in revealing candidate SNPs. Moreover, from literature review, Lin et. al. (2012) [15] and Lasheras et. al. (2017) [14] both combined Random Forest and Deep Learning-Convolution Neural Network (CNN) respectively, with MARS. They showed that by combining another method with MARS, the resulting MARS model greatly improved than when MARS was used alone. Therefore, I am likewise investigating how to improve the MARS model by combining it with another method. Thus, I am currently working on combining the Random Forest (RF), embeded in **Recursive Feature Elimination (rfe/ RFE)** method with MARS, for the next phase of this study. RFE is a variable selection machine learning method that works to select variables by recursively considering smaller and smaller sets of variables. Here is a little detail of how RFE works: Initially, the estimator is trained on the first set of variables (features) and the importance of each variable is achieved through a feature-importance attribute. Next, the least important variables are removed from the current set of variables. This process is recursively repeated on the pruned set until the desired number of variables required to be selected is eventually obtained.

In the Python Programming Language, the package Scikit Learn is able to easily implement RFE by passing any classifier through it. For this study, the classifier used is RF. Originally, the planned was to use PCA (Did a mini project on it already. Ultimately, rfe(RF) was settled on due to its relatively effectiveness and ease of interpretation.

The table below (figure 4 display some preliminary results of rfe(RF) plus MARS. The recovery rate of the known SNPs is 100 percent for datasets one and two. My next plan of action along this line will be:

- To Simulate two more datasets of width 100 and 1000 SNPS respectively with known SNPs:
- Then, apply rfe(RF) plus MARS to them:
- Finally, I will apply rfe(RF) plus MARS to the main data or any real SNP data.

## 6 Conclusions

The three methods for the study of gene-gene interactions we identified are MDR, RF + logistic regression, and MARS. Comparing the three methods, MDR was the most accurate in the test set (though was extremely slow in case of large data size) since unlike the others, it’s recovery rate was 100%. However, MARS was faster in revealing candidate SNPs in the chronicity DILI data set. On the chronicity DILI dataset, MARS retrieved the SNP, rs6487213, as part of its interacting SNPs. We identified this SNP as a suspected allelic variation of the gene SLCO1B1. This gene is a known predictor of elevated serum bilirubin, which is a risk factor related to chronic DILI [11].

Though I just started applying the rfe(RF) plus MARS method to some of the data, so far it seems promising and the recovery rate has been 100 percent for datasets one and two.

### 6.1 Ongoing Work

- I will continue the work on the rfe(RF) plus MARS method and complete it. I may also later combine MARS with a Deep Learning method e.g. Convolutional Neural Network.

### 6.2 Future Work

In addition to developing MARS with other methods for DILI, we plan to later apply/extend found methods to genetic DILI data with covariates/ environmental factors.

## Data Availability

All data used are readily available on the website links provided next.

https://ani.stat.fsu.edu/~vic/DILIdata/

## 6.3 Acknowledgement

I am especially grateful to my advisor, Dr. Victor Patrangenaru, who apart from being a great advisor and professor to me, allowed me to continue the DILI research from my Summer 2018 Internship, and patiently guided me through this whole PhD essay with invaluable suggestions and editing.

## 6.4 Funding

This research received no external funding.

## 6.5 Conflicts Of Interest

The authors declare no conflict of interest.

## Notes

### Competing Interest Statement

The authors have declared no competing interest.

### Clinical Trial

This is not a clinical trial.

### Funding Statement

No external funding was received

### Summary of Updates

Edited authors' degree information. Also, some unnecessary details for a few sections were removed, but the main concept and information remained unchanged.

## References

[1] Agresti, A. (2018). An introduction to categorical data analysis. Hoboken, NJ: John Wiley & Sons.

[2] Breiman, L. (2001). Random forests. Machine learning, 45(1), 5–32.

[3] Chen, M., Suzuki, A., Borlak, J., Andrade, R. J., & Lucena, M. I. (2015). Drug-induced liver injury: Interactions between drug properties and host factors. Journal of Hepatology, 63(2), 503–514. doi:10.1016/j.jhep.2015.04.016

[4] Cutler, Adele & Cutler, David & Stevens, John. (2011). Random Forests. 10.1007/978-1-4419-9326-75.

[5] Daly AK, Donaldson PT, Bhatnagar P, et al. HLA-B*5701 genotype is a major determinant of drug-induced liver injury due to flucloxacillin. Nat Genet. 2009;41(7):816–819

[6] Friedman, J. H. (1991). Multivariate Adaptive Regression Splines. The Annals of Statistics, 19, 1–67.

[7] Gola, D., John, J. M. M., Steen, K. V., & König, I. R. (2015). A roadmap to multifactor dimensionality reduction methods. Briefings in Bioinformatics, 17(2), 293–308. doi: 10.1093/bib/bbv038

[8] Goldstein, Benjamin A, et al. (2011) Random Forests for Genetic Association Studies. Statistical Applications in Genetics and Molecular Biology, vol. 10, no. 1, Dec. 2011, doi:10.2202/1544-6115.1691.

[9] Home: iSAEC. (n.d.). Retrieved January 1(2019), from http://www.saeconsortium.org/

[10] Human Genome Project Information. (1999, January 1). Retrieved July 1, 2017, from http://www.herbogeminis.com/IMG/pdf/snp.pdf

[11] Johnson, A. D., Kavousi, M., Smith, A. V., Chen, M., Dehghan, A., Aspelund, T., Witteman, J. C. (2009). Genome-wide association meta-analysis for total serum bilirubin levels. Human Molecular Genetics, 18(14), 2700–2710. doi:10.1093/hmg/ddp202

[12] Kane, D. (2015, June 30). Data Science - Part XV - MARS, Logistic Regression, & Survival Analysis. Retrieved from https://www.youtube.com/watch?v=17QbQF9XM&t=1876s

[13] Kaplowitz, N. (2013). Drug-induced liver injury: introduction and overview. In Drug-Induced Liver Disease (pp. 3–14). Academic Press.

[14] Lasheras, J. E. S., Tardón, A., Tardón, G. G., Gómez, S. L. S., Sánchez, V. M., Donquiles, C. G., & de Cos Juez, F. J. (2017, September). A methodology for the detection of relevant single nucleotide polymorphism in prostate cancer by means of multivariate adaptive regression splines and back-propagation artificial neural networks. In International Joint Conference SOCO2017-CISIS2017-ICEUTE2017 León, Spain, September 6–8, 2017, Proceeding (pp. 391–399). Springer, Cham.

[15] Lin, H. Y., Ann Chen, Y., Tsai, Y. Y., Qu, X., Tseng, T. S., & Park, J. Y. (2012). TRM: A Powerful Two-Stage Machine Learning Approach for Identifying SNP-SNP Interactions. Annals of human genetics, 76(1), 53–62.

[16] Lou, Xiang-Yang, et al. “A Generalized Combinatorial Approach for Detecting Gene-by-Gene and Gene-by-Environment Interactions with Application to Nicotine Dependence.” American Journal of Human Genetics, The American Society of Human Genetics, June 2007, www.ncbi.nlm.nih.gov/pmc/articles/PMC1867100/.

[17] Medina-Caliz, I., Robles-Diaz, M., Garcia-Muñoz, B., Stephens, C., Ortega-Alonso, A., Garcia-Cortes, M., … Andrade, R. J. (2016). Definition and risk factors for chronicity following acute idiosyncratic drug-induced liver injury. Journal of Hepatology, 65(3), 532–542. doi:10.1016/j.jhep.2016.05.003

[18] Moore, J. H. (2010). Detecting, Characterizing, and Interpreting Nonlinear Gene–Gene Interactions Using Multifactor Dimensionality Reduction. Computational Methods for Genetics of Complex Traits Advances in Genetics, 101–116. doi:10.1016/b978-0-12-380862-2.00005-9

[19] Moore, J. H. and Andrews, P. C. (2015). Epistasis Analysis Using Multifactor Dimensionality Reduction. Methods in Molecular Biology Epistasis, 301–314. doi: 10.1007/978-1-4939-2155-316

[20] Niel, C., Sinoquet, C., Dina, C., & Rocheleau, G. (2015). A survey about methods dedicated to epistasis detection. Frontiers in Genetics, 6. doi: 10.3389/fgene.2015.00285

[21] Ritchie, M. D. and Motsinger, A. A. (2005). Multifactor dimensionality reduction for detecting gene–gene and gene–environment interactions in pharmacogenomics studies. Pharmacogenomics, 6(8), 823–834. doi:10.2217/14622416.6.8.823

[22] Salford Systems. ”Multivariate Adaptive Regression Splines (MARS)” Random Forests OOB vs. Test Partition Performance - Dan Steinberg’s Blog, www.salford-systems.com/products/mars

[23] Shang, J., Zhang, J., Sun, Y., Liu, D., Ye, D., & Yin, Y. (2011). Performance analysis of novel methods for detecting epistasis. BMC Bioinformatics, 12(1). doi: 10.1186/1471-2105-12-475

[24] Urban, T. J.; Goldstein, D. B.; Watkins, P. B. (2012). Genetic basis of susceptibility to drug-induced liver injury: What have we learned and where do we go from here? Pharmacogenomics, 13(7), 735–738. doi:10.2217/pgs.12.45

[25] Wei, W.-H., Hemani, G., & Haley, C. S. (2014). Detecting epistasis in human complex traits. Nature Reviews Genetics, 15(11), 722–733. doi: 10.1038/nrg3747

[26] www.statsoft.com. (n.d.). Retrieved from http://www.statsoft.com/Textbook/Multivariate-Adaptive-Regression-Splines

[27] Xu, H., Sun, X., Qi, T., Lin, W., Liu, N., & Lou, X. (2014). Multivariate Dimensionality Reduction Approaches to Identify Gene-Gene and Gene-Environment Interactions Underlying Multiple Complex Traits. PLoS ONE, 9(9). doi:10.1371/journal.pone.0108103

[28] Yang, C.-C., Prasher, S. O., Lacroix, R., & Kim, S. H. (2004). Application Of Multivariate Adaptive Regression Splines (Mars) To Simulate Soil Temperature. Transactions of the ASAE, 47(3), 881–887. doi: 10.13031/2013.16085

[29] Yi, N. (2010). Statistical analysis of genetic interactions. Genetics Research, 92(5-6), 443–459. doi: 10.1017/s0016672310000595

[30] York, Timothy P., and Lindon J. Eaves. (2001). Common Disease Analysis Using Multivariate Adaptive Regression Splines (MARS): Genetic Analysis Workshop 12 Simulated Sequence Data. Genetic Epidemiology, vol. 21, no. S1, 2001, doi:10.1002/gepi.2001.21.s1.s649.

